# Pharmaco-nutritional strategies to increase nitric oxide signaling in Raynaud’s phenomenon (Nivose): a series of N-of-1 trials

**DOI:** 10.64898/2026.08.13.26360355

**Authors:** Alicia Guigui, Marc Manceau, Joris Giai, Clément Jam-bon-Barbara, Adeline Paris, Jean-Luc Cracowski, Matthieu Roustit, Charles Khouri

**Affiliations:** Univ. Grenoble Alpes, Inserm U1300, HP2 Laboratory, Grenoble, France; Univ. Grenoble Alpes, Inserm CIC1406, CHU de Grenoble, Grenoble, France; Univ. Grenoble Alpes, Inserm, CHU Grenoble Alpes, CIC 1406, 38000, Grenoble, France; Univ. Grenoble Alpes, CNRS, UMR 5525, VetAgro Sup, Grenoble INP, TIMC, 38000 Grenoble, France; Univ. Grenoble Alpes, Pharmacovigilance Unit, Grenoble Alpes University Hospital, F-38000 Grenoble, France

## Abstract

**Background:** Treatment of Raynaud phenomenon(RP) with oral vasodilators(calcium channel blockers and phosphodiesterase type 5 inhibitors) has shown moderate efficacy, may not benefit to all patients, and adverse effects often compromise long-term treatment. In addition, a large placebo effect may jeopardize the assessment of treatment benefits. Pharmaconutritional strategies aiming at increasing nitric oxide bioavailability (beetroot juice and L-citrulline) may be promising alternatives, and we further hypothesized that patient preference for a treatment could be a driver of the response.

**Methods:** This study consisted of a series of randomized, double-blind, N-of-1 trials conducted in outpatients with primary or secondary RP. Each patient underwent a multiple crossover design with repeated blocks of randomized treatments periods: 2 weeks of placebo, 2 weeks of active treatments, and 1 week of washout. Outcomes included the Raynaud Condition Score(RCS), frequency and daily duration of attacks. Each patient prespecified its preferred primary outcome, efficacy threshold and preferred treatment, which was used for stratified randomization. Generalized linear mixed-effects models were used to determine individual and aggregated efficacy.

**Results:** Twenty-one patients completed 2 to 8 treatment blocks. Seventeen patients tested L-citrulline, 17 beetroot juice and 13 both treatments. Ten patients selected RCS as a primary outcome, 6 patients the number of attacks and 5 the duration of attacks. Median threshold for considering treatment efficacy chosen by patients was 50% (min-max 20% to 75%) reduction of symptoms. Using individual criteria to define efficacy neither L-citrulline nor beetroot juice showed significant efficacy compared to baseline. Based on the aggregated data, our results show no significant difference between L-citrulline and the L-citrulline-based placebo, nor between beetroot juice and nitrate-depleted beetroot juice, with the exception of the daily duration of RP attacks with beetroot juice (p=0.002). Finally, there was a marked placebo response, notably when patients received their preferred treatment.

**Conclusions:** Our study did not show significant beetroot juice or L-citrulline efficacy in RP. However, we found that individual preference for one treatment over another maximizes responses to both placebo and active treatments, particularly with regard to the frequency and duration of RP attacks, thus suggesting that a real and modifiable placebo effect exists in RP.

**Trial registration:** NCT03749577

**Funding:** Primary Funding Source: Grenoble Alpes University Hospital & Association des Sclérodermiques de France (academic funding).

## Introduction

Raynaud’s phenomenon (RP) is a condition characterized by biphasic or triphasic skin color changes (pallor, cyanosis and the hyperemia of reflow) of the fingers or toes triggered by cold or emotional stress [1]. Primary RP is relatively common, affecting 2-12% of the general population. It is generally a seasonal and benign disease, responding to conservative measures, such as avoiding exposure to cold and wearing protective clothing [2]. Secondary RP is associated with a variety of conditions (particularly autoimmune connective tissue disease such as systemic sclerosis, SSc), exposures and drugs [3]. When severe, RP can progress to digital ulceration and/or critical digital ischemia. The pathophysiology of RP remains unclear, but microvascular dysfunction and alteration of the NO-dependent endothelial vasodilation pathway via NO synthases (NOS) represent one of the main mechanisms [1].

A pharmacological treatment in RP is proposed to patients when conservative measures are not efficient enough to control the symptoms [4]. General lifestyle measures (smoking cessation, cold exclusion and keeping warm) are always recommended. If these are insufficient, extended-release calcium channel blockers (CCB) remain the first line pharmacological treatment of RP. When CCBs fail to adequately control symptoms, cause adverse effects or when RP is complicated by digital ulceration, phosphodiesterase type 5 inhibitors (PDE5i) may be initiated or added. Lastly, intravenous iloprost is considered for severe RP following failure of oral therapy [5,6]. Yet, these drugs are commonly associated with adverse effects that often result in permanent discontinuation, and they have shown very modest efficacy compared with placebo in clinical trials in primary and secondary RP [7,8]. Thus, patients may prefer complementary and alternative therapies, even though none has proven its efficacy.

In this trial we tested two pharmaco-nutritional strategies aiming at increasing nitric oxide (NO) bioavailability through L-citrulline and beetroot juice administration. L-citrulline is indeed transported into endothelial cells and then converted into L-arginine, a substrate for NO synthase [9,10]. Previous studies have shown efficacy of an oral supplementation of L-citrulline to increase NO synthesis [11,12]. Beetroot juice is an important source of inorganic nitrates, which are converted to nitrites through the enterosalivary pathway, and have been successfully used to increase NO bioavailability, to reduce blood pressure and to improve endothelium-dependent and -independent vasodilation in the forearm [13,14].

In this series of single patient trials (N-of-1), we are testing the hypothesis that L-citrulline or beetroot juice are efficient to decrease the severity of RP, compared to their respective placebo. In addition, considering the large placebo response in Raynaud trials [15], we further hypothesized that patient preference could be a driver of the response. We thus asked patients to define their preferred treatment strategy before the trial started, and stratified randomization accordingly. As such, in this pragmatic patient-centered approach, we let patients choose their primary outcome and the minimal clinically relevant difference used as a personal threshold of significant efficacy (referred to as “individual efficacy margin” in the rest of the manuscript).

## Methods

### 1. Design of the trials

This study followed the CONSORT (Consolidated Standards of Reporting Trials) CENT (extension for reporting N-of-1 trials) 2015 Statement [16]. NIvoSe is a single center study performed at the Clinical Research Center of the Grenoble Alps University Hospital.

This study consists of a series of N-of-1 trials. Each N-of-1 trial is a prospectively planned, multiple crossover study in a single individual. This design is particularly suited to assess treatments for RP, as heterogeneity in daily activities, in environmental conditions, as well as in the severity of RP induces significant variability and makes efficacy difficult to demonstrate in usual randomized controlled trials (RCT) [17].

The study was conducted over three winter seasons, each participant being proposed to participate for two consecutive winters. For each of them, a two-weeks run-in period was first conducted to assess the frequency, duration and severity of RP at baseline, and for the patient to become familiar with recording daily symptoms in a paper diary. The treatment proposed to patients in the first winter (L-citrulline or beetroot juice) was randomized and stratified on the patient preference (L-citrulline, beetroot or no preference, see below). Then all patients were proposed to test the alternative intervention during the next winter season.

Overall, two to six blocks were conducted over two consecutive winters, one block consisted of two 2-weeks supplementation periods:

- L-citrulline 9.6g/day or beetroot juice 70ml/day and
- L-citrulline placebo (maltodextrin) or beetroot juice placebo (nitrate-depleted beetroot juice).

In order to exclude any carryover effect, a 7 days wash-out period was imposed between treatment periods. The sequence, i.e. the order of periods within each block, was also randomized. Both investigators and patients were blinded. **Figure 1A** summarizes the experimental plan.

**Figure 1.**
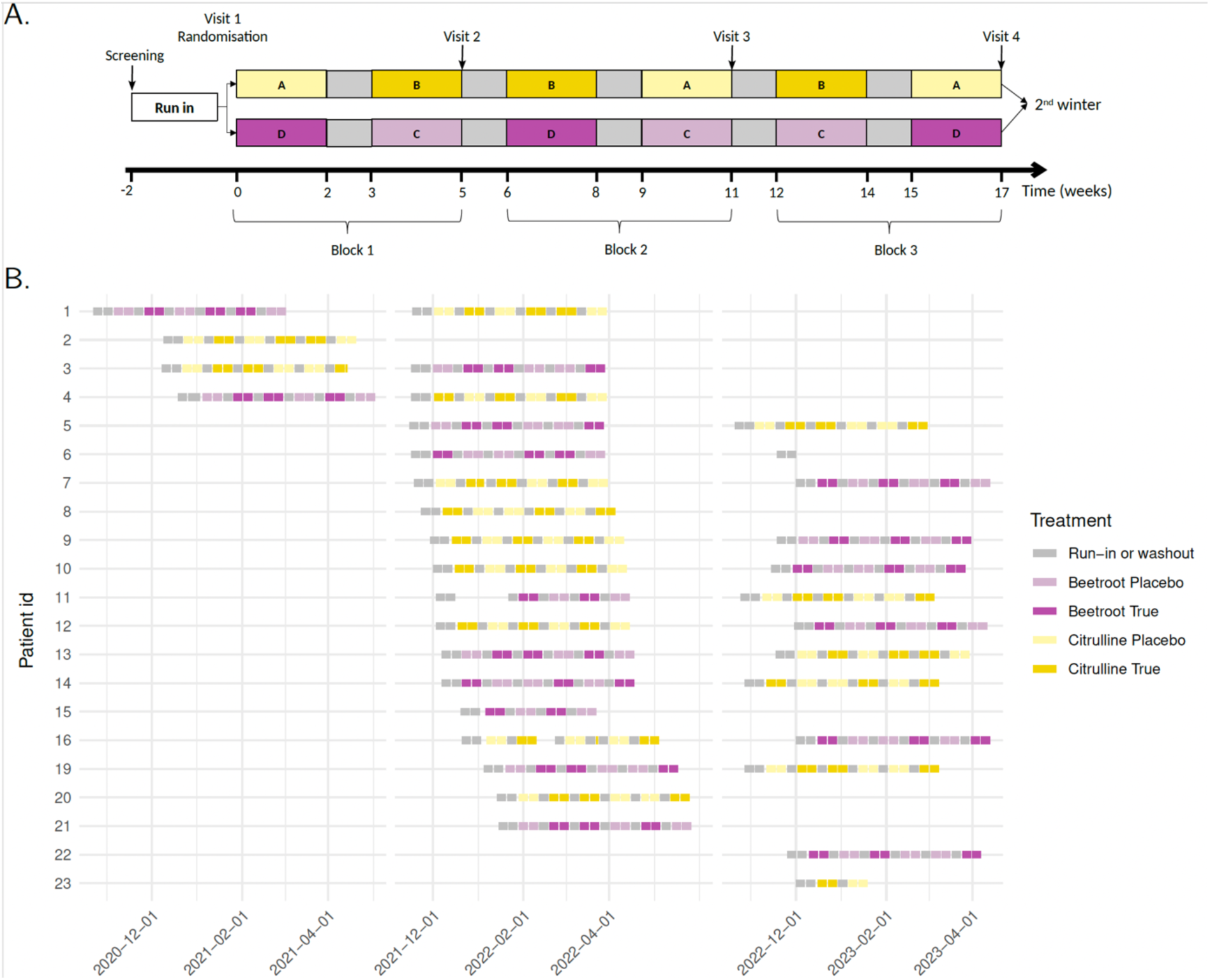
A. Study design including 3 blocks of 3 periods with a crossover in the following winter. Patients are randomized in arm 1 or 2 after a run-in period to test beetroot juice or L-citrulline and are asked to test the alternative treatment arm in a second winter. The order of treatments within each block is randomized. B. Precise timeline of the N-of-1 trials for the 21 patients included.

### 2. Participants

The investigation conformed to the principles outlined in the Declaration of Helsinki. The protocol of the study was approved by the Ethics Committee *Comité de Protection des Personnes Est II* (Institutional Review Board 18/591) and the National Agency for Medicines Safety (ANSM), and each patient gave written informed consent before participation. Patients were recruited through the vascular medicine department of Grenoble Alps University Hospital and enrolled at the clinical pharmacology unit between December 2020 and April 2023. All participants were at least 18 years of age, had a Body Mass Index>18, and had primary or secondary active RP, with a stable disease over the previous two months. Active RP was characterized by a clinical history of Raynaud’s: 1/ Primary RP or secondary to limited (LSSc), limited cutaneous (lcSSc) or diffuse cutaneous scleroderma dcSSc) according to the criteria of Leroy and Medsger [18]; 2/ At least 1 RP attacks per week and a specific finger patient picture in RP attack (assessed over the 2 weeks preceding inclusion). The main exclusion criteria are listed in **Supplement Table 1.** For women of childbearing age, a contraceptive method was needed (hormonal, physical or abstinence).

### 3. Intervention

The nutritional strategies that have been tested are:

– L-citrulline 9.6g per day (capsule, per os) and L-citrulline placebo (maltodextrin) (capsule, per os) during 14 days; purchased from NOW FOODS®. Both treatments were blinded by the Pharmacy department, Grenoble Alps University Hospital.
– Concentrated 70 ml beetroot juice bottles containing 6.45 mmol of nitrates (Beet It®), and nitrate-depleted beetroot juice were purchased from James White Drinks, Ashbocking, UK. Both treatments were blinded by the Pharmacy department, Grenoble Alps University Hospital.

### 4. Outcomes

Principal outcomes were the Raynaud Condition Score (RCS), the frequency, and cumulative duration of attacks over 24 hours. After information about these outcomes, each volunteer chose his/her own main outcome among these 3 criteria (the two non-chosen outcomes were used as secondary endpoints, see below), as well as its associated individual efficacy margin, corresponding to the minimum expected benefit for the treatment to be worthwhile (e.g. decreasing the frequency of attacks from 30%). Outcomes were collected by using daily diary cards. When an attack occurred, patients recorded the time the event started and ended in the diary, and its duration was subsequently calculated, as previously described [17]. Moreover, patients were asked to daily fulfill the Raynaud Condition Score (RCS) which has been previously translated and validated in French [19]. Outcomes were collected during run-in, treatment and washout periods.

The secondary objectives were:

1/ To compare the aggregated efficacy of a 2-week supplementation with L-citrulline or beetroot juice versus their respective placebos, on all clinical outcomes (RCS, daily frequency or duration).
2/ To compare the safety of a 2-week supplementation with L-citrulline and beetroot juice versus their respective placebos, assessed through adverse events collected in the daily dairies’ cards. Adverse events were graded according to Common Terminology Criteria for Adverse Events, version 4.0.
3/ To assess the placebo response of L-citrulline placebo and nitrate-depleted beetroot juice compared to run-in and washout periods, and according to patient preference.

### 5. Sample size calculation and statistical analysis

In N-of-1 trials, there is properly speaking no sample size calculation for the estimation of individual effects since the precision depends on the number of blocks within a single participant.

We analyzed the three outcomes of interest within a generalized linear mixed model (GLMM) framework, with a random intercept for each patient and fixed effects corresponding to the following covariates: treatment received, gender, age at inclusion, primary or secondary Raynaud’s phenomenon, the minimal temperature on that day and cumulative rainfall on that day. Daily mean temperature and humidity from the weather station closest to each patient’s home were collected to assess the influence of environmental factors on the onset of attacks.

The individual baseline was calculated for each outcome as an average of the run-in and washout periods to avoid regression toward the mean phenomenon.

More precisely, the distributions considered in our GLMM were the following. First, the number of attacks was assumed to be Poisson distributed with a rate parameter depending on all these covariates. Second, the cumulative duration of attacks was assumed to be Gamma distributed with the shape parameter being the number of attacks and the rate parameter depending on all covariates. This corresponds to a model where each attack has an exponentially distributed duration, thus considering the expected correlation between the number of attacks and the cumulative duration of attacks. Third, the RCS is an integer between 0 and 10, which is modeled using a binomial distribution with a fixed number of trials (10) and a probability of getting each point of the scale depending on all covariates.

For the primary objective, each patient is classified as responder under each treatment when the difference between his/her expected preferred outcome under the treatment and his/her expected preferred outcome during wash-out and run-in periods is greater than his/her individual efficacy margin.

For the secondary objectives:

1/ The fixed effects of L-citrulline and L-citrulline placebo or the fixed effects of beetroot juice or nitrate-depleted beetroot juice, were compared. Given the number of tests performed we used a Bonferroni correction of the p-value; and considered significant p-values <0.0083.
2/ The safety comparison of the proportion of adverse events under the four treatment conditions was performed using a chi square test.
3/ The fixed effects of L-citrulline placebo and nitrate-depleted beetroot juice were compared to zero. A second version of the model was also fitted to the data, including a new covariate indicating whether the preference of the patient and the treatment were aligned, and the associated fixed effect is compared to zero as well. Given the number of tests performed we used a Bonferroni correction of the p-value; and considered significant p-values<0.017.

## Results

### 1. Participants

Twenty-three individuals were enrolled in this study, two patients did not complete the study and were excluded from the analysis (**Figure 2**). Twenty-one patients completed the study, with 17 testing L-citrulline, 17 testing beetroot juice, and 13 testing both treatments. Patients completed a mean of 6.33 (SD 2.15) treatment periods corresponding to a mean of 207.52 (SD 74.22) days of observation (**Supplementary Figure 1 & Figure 1B**).

**Figure 2.**
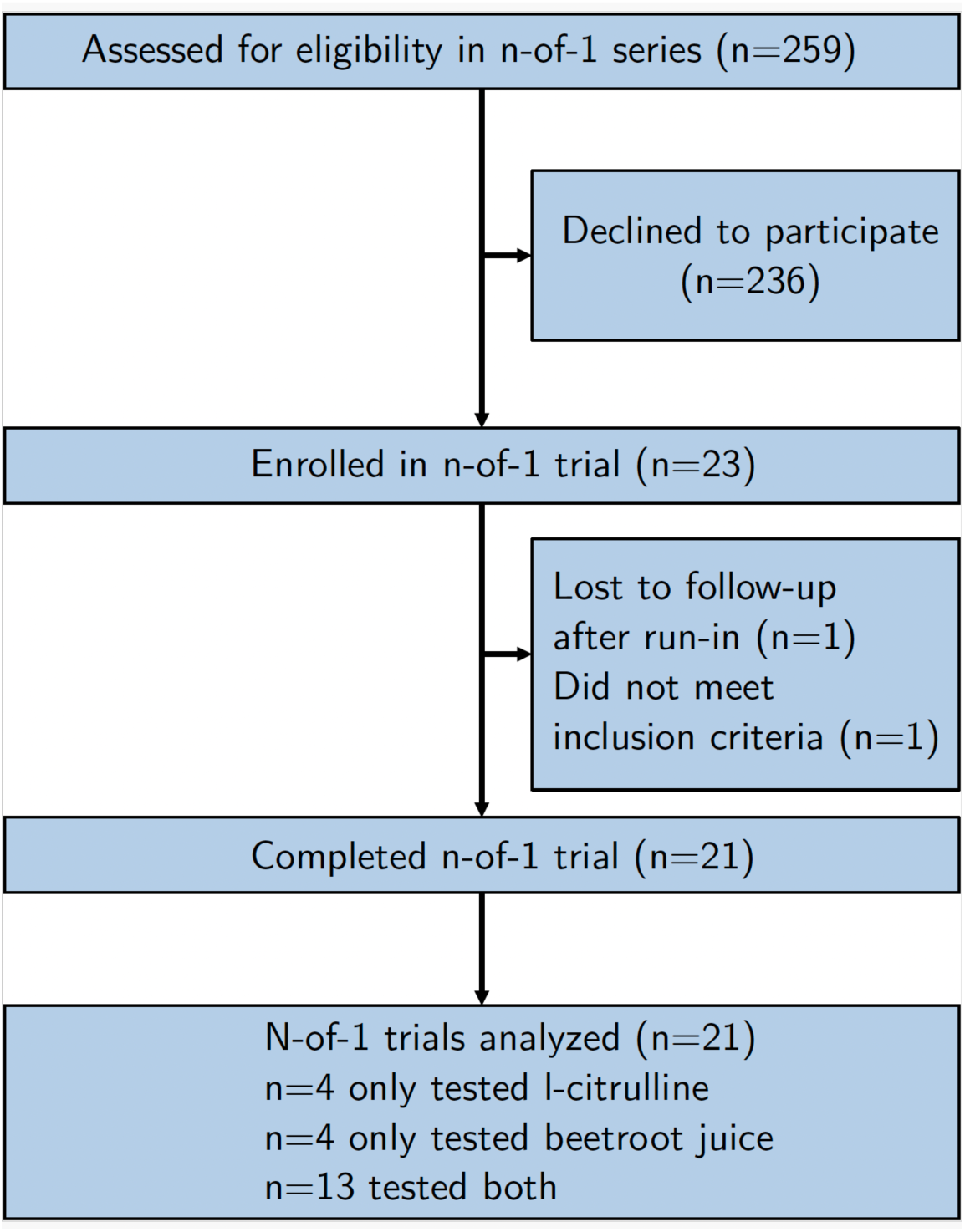
CONSORT CENT Flow diagram.

Characteristics of patients are presented in **Table 1**. Most patients were female with primary RP, and only 1 patient was treated with CCB at baseline.

**Table 1.** Characteristics of the 21 included patients.

| Characteristic | Value |
| --- | --- |
| Mean age (min-max) | 43 (35 - 59) |
| Female, n (%) | 18 (85.7%) |
| Cause of RP, n (%) |  |
| Primary | 19 (90.5%) |
| Secondary | 2 (9.5%) |
| Number of fingers involved |  |
| 6 | 3 (15%) |
| 8 | 13 (65%) |
| 10 | 4 (20%) |
| Smoking |  |
| Active smoker | 2 (10.5%) |
| Former | 5 (26.3%) |
| Never smoked | 12 (63.2%) |
| Oral vasodilators | 1 (4.8%) |

During run-in periods, patients presented a median of 1.14 (Q1-Q3 0.89-1.59) crisis per day with a cumulative duration of 39.71 (Q1-Q3 23.20-72.33) minutes per day and a RCS of 3.43 (Q1-Q3 2.57-5.64) points.

### 2. Choice of outcomes, threshold and patient preference

Ten patients selected RCS as a primary outcome, 6 patients the number of attacks and 5 the duration of attacks. Median threshold for considering treatment efficacy chosen by patients was 50% (min-max 20% to 75%) reduction of symptoms. Considering their baseline severity, patients having chosen RCS as a primary outcome expected a reduction of 1.75 (Q1-Q3 1.05-2.05) points. In patients having chosen the frequency of attacks it was 0.64 (Q1-Q3 0.50; 0.65) attacks per day and 22.40 (Q1-Q3 7.54-31.16) minutes per day for patients having chosen the duration of attacks. Individual description of patient’s choices and baseline severity of RP is provided in **Supplementary Table 2.**

### 3. Individual and aggregated treatment efficacy

#### Individual treatment efficacy

The individual percentage of reduction of adjusted RCS, number of attacks/day and cumulative daily duration of RP are presented in **Figure 3**. Based on individual efficacy margins and patient-defined primary outcomes, neither beetroot juice nor L-citrulline have a significant effect in any patient.

**Figure 3.**
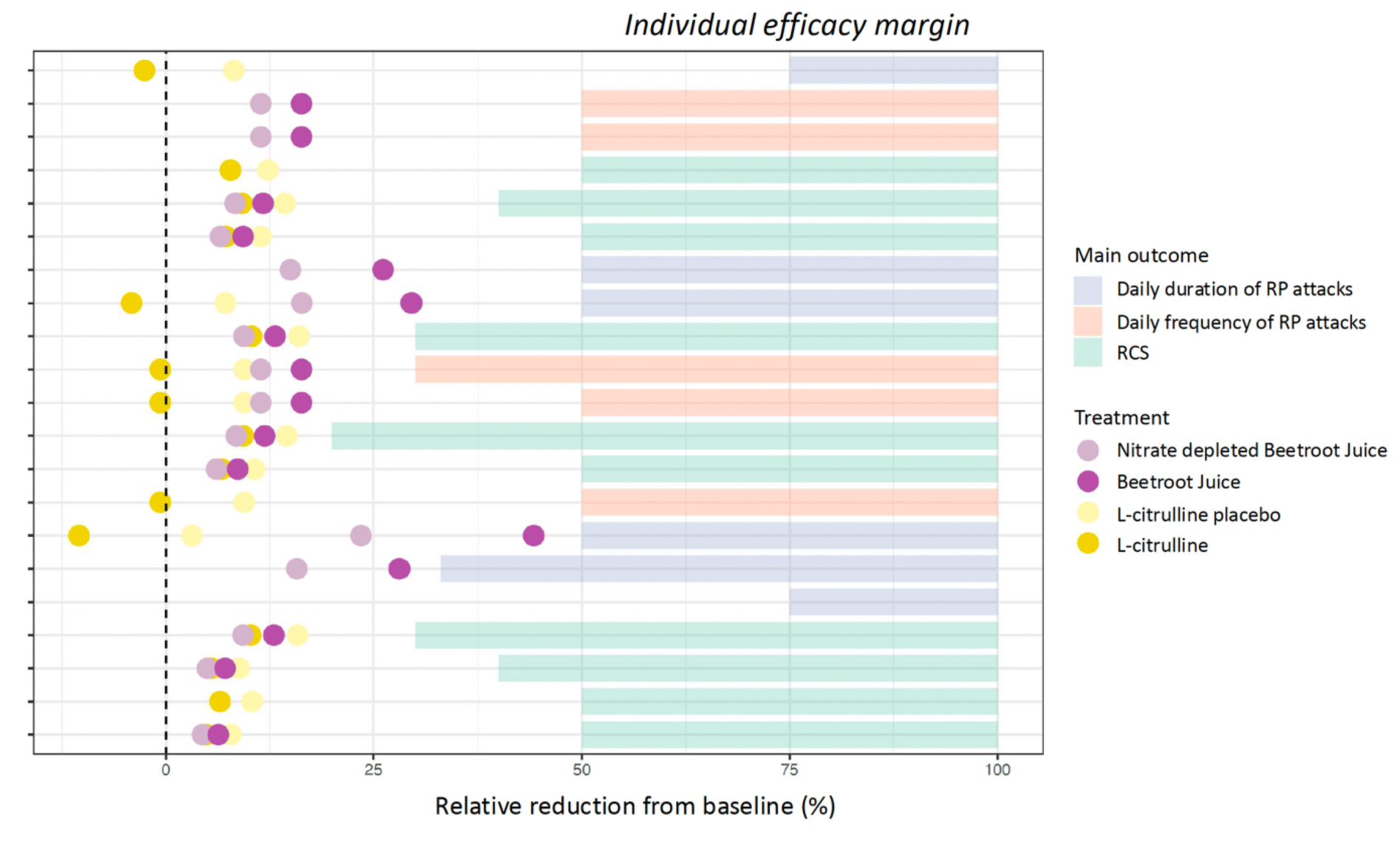
Predicted individual responses under the different treatments tested.

#### Aggregated treatment efficacy

The comparison of the effect of L-citrulline and L-citrulline placebo is non-significant on all outcomes. The comparison of the effect of beetroot juice and nitrate-depleted beetroot juice is non-significant on RCS and on the number of attacks/days, but is significant on the cumulative duration of attacks (p=0.002). Adjusted outcomes of all individuals included in the study under different treatments are presented in **Figure 4**.

**Figure 4.**
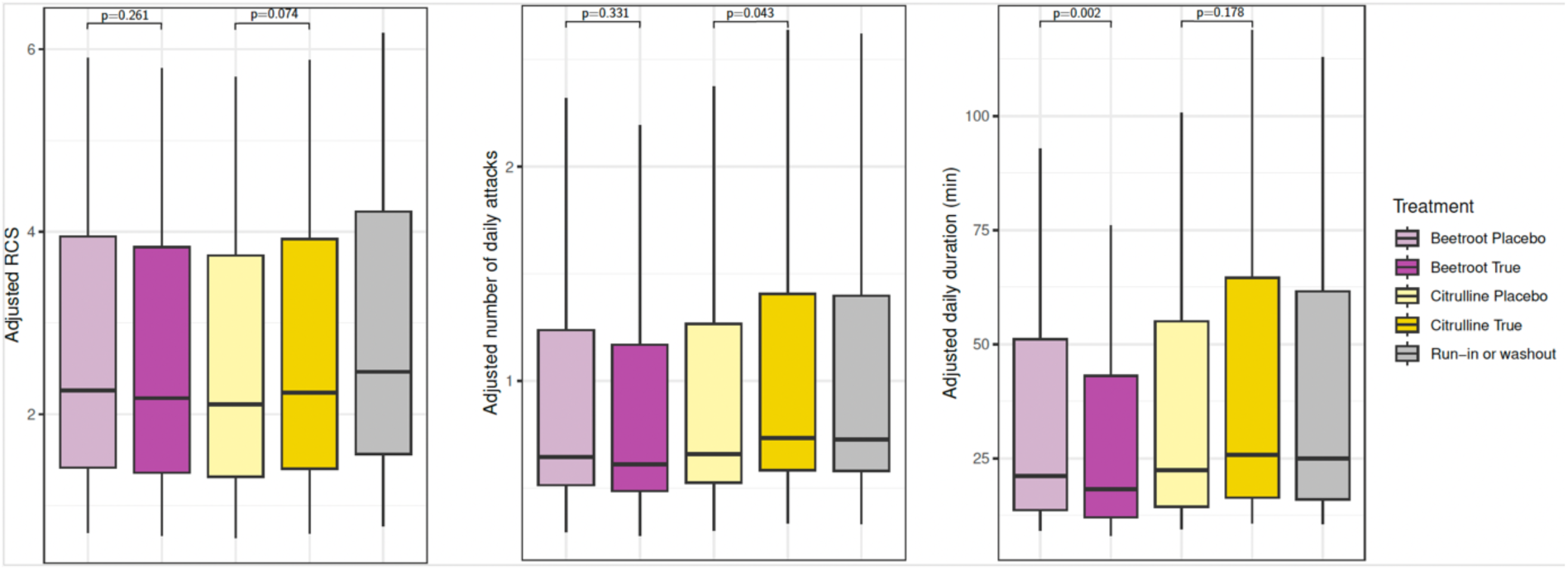
Adjusted outcome predicted under different conditions among all individuals included in the study to assess the overall placebo response.

### 4. Safety

A total of 66 adverse events (AEs) were reported during the study, none of which were classified as serious. There was no significant difference in AE incidence between active treatments and their respective placebos, or across treatment arms (p = 0.35). Under beetroot juice supplementation, most AEs were mild and included headaches (n = 2), presyncope, diarrhea (n = 2), nasopharyngitis (n = 2), oropharyngeal pain, neck pain, back pain, and influenza-like illness. Two cases of COVID-19 were also reported. During L-citrulline supplementation, AEs were infrequent and included viral pharyngitis, diarrhea, two cases of influenza-like illness, and one case of COVID-19. Only diarrhea was considered related to the study intervention.

### 5. Relationship between treatment preference and placebo response

We found a significant interaction between patient preference for a treatment and placebo response (i.e. patients randomized on their preferred treatment displayed higher treatment response, whether active or placebo, than patient randomized in the other treatment). Interestingly, this interaction was significant for the frequency (p=0.0008) and duration (p<0.0001) of attacks but not for RCS (p=0.051; adjusted threshold at 0.017) **(Figure 5)**. Despite the absence of inclusion criteria based on RP severity, squeeze plots comparing run-in with no treatment or washout periods suggest a regression toward the mean phenomenon (**Supplementary Figure 2**).

**Figure 5.**
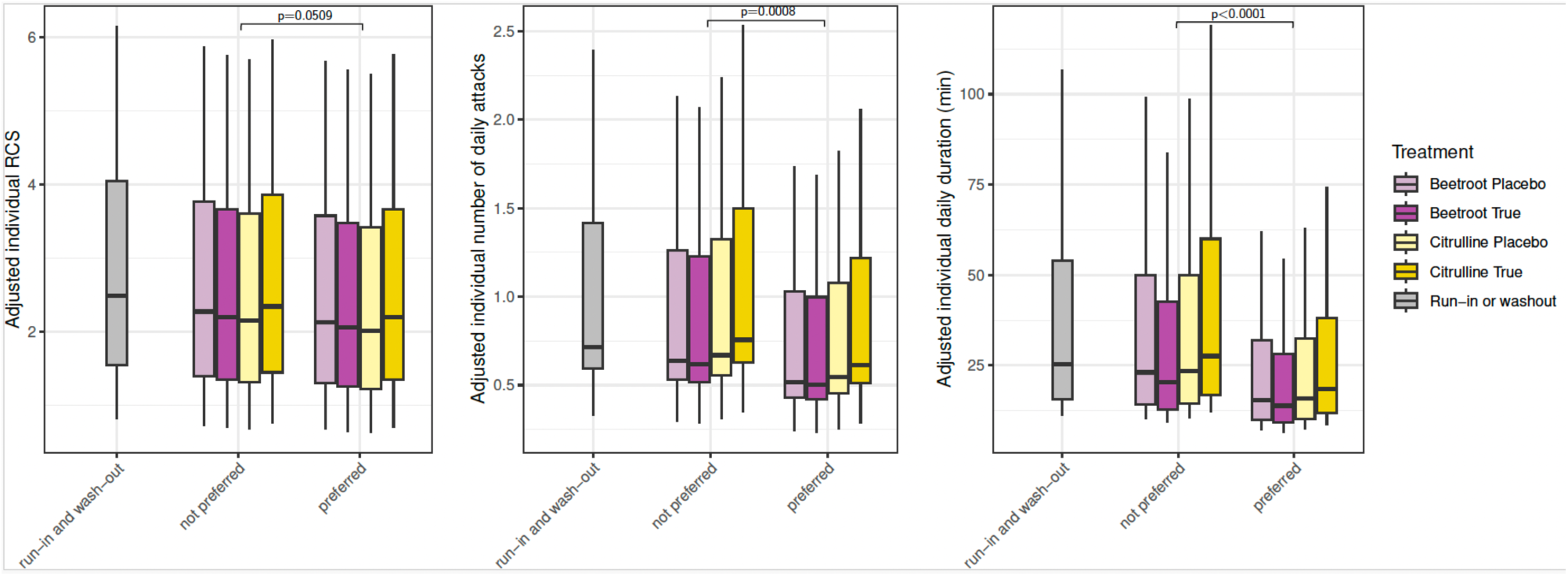
Adjusted outcome predicted under different conditions among all individuals included in the study to assess the overall effect of getting the preferred treatment.

## Discussion

In this series of N-of-1 trials, involving 21 patients, we found that, compared to baseline, neither L-citrulline nor beetroot juice improved RP according to the criteria and margins defined by the patients themselves. Based on the aggregated data, our results showed no significant difference between L-citrulline and the L-citrulline-based placebo, nor between beetroot juice and nitrate-depleted beetroot juice, with the exception of the daily duration of RP attacks with beetroot juice. However, we found that the response to treatments, whether active or placebos, was significantly higher on the frequency and duration of RP attacks in patients who actually received their preferred treatment.

The rationale for testing L-citrulline and beetroot juice was rooted in several pre-clinical and clinical studies. Indeed, both treatments have demonstrated their ability to increase NO bioavailability [20–23]. A previous study notably showed that both beetroot juice and nitrate-depleted beetroot juice increased blood flow in the thumb following a cold challenge; improved endothelium-dependent and -independent vasodilation in the forearm; and inflammatory markers [13]. Similarly, compared to baseline values, we observed a reduction in all RP-related outcomes during periods when patients were given beetroot juice, but also nitrate-depleted beetroot juice (not tested). In addition, we observed a potential effect of beetroot juice on the duration of attacks compared to nitrate-depleted beetroot juice. It is worth noting that, in a previous N-of-1 trial testing on-demand sildenafil, we also found a greater effect of sildenafil on this outcome [17]. We can therefore hypothesize that it could be mediated by NO bioavailability and explained by faster reperfusion of the skin microvasculature after cold exposure; such an effect has also been observed in healthy volunteers after ingesting beetroot juice [24].

Surprisingly, these results were not replicated with L-citrulline. One possible explanation could be that L-citrulline requires functional NO synthases to be converted into NO, and recent studies have shown a decrease in endothelial NO synthase expression in patients with RP [25].

The use of N-of-1 trials is particularly suited to treatment evaluation in RP, a highly variable condition, as it increases statistical power by controlling for patient-specific covariates, and allows to daily adjust for meteorological conditions. This improves the accuracy of aggregate estimates and reduces sample size requirements. Beyond methodological considerations, it is a patient-centered approach that allows estimation of individual treatment effects. In this study, we further asked patients to choose their primary efficacy endpoint with its associated efficacy margin.

The diversity of patient-prioritized endpoints was unexpected, as they were distributed across all three commonly used outcomes. Definition of endpoints in clinical trials in RP has long been debated and echoes recent development of patient-reported outcomes for systemic sclerosis associated RP [26–28]. Our results also reinforce that the RCS represents a broader dimension of the disease than other endpoints, with patients reporting a non-zero RCS even though they did not actually experience any PR attacks during the day even in primary RP patients [29].

Moreover, efficacy margins chosen by patients reflect the high expectation of patients for a treatment effect, in light of the modest effectiveness of available therapies. When estimating the individual minimal clinically important differences using the severity of the disease in the run-in period and the individual efficacy margin, the results translate in a reduction of 1.75 (Q1-Q3 1.05-2.05) point of RCS, 0.64 (Q1-Q3 0.50; 0.65) attacks per day and 22.40 (Q1-Q3 7.54-31.16) minutes per day. Interestingly, the result for the RCS is coherent with the defined minimal clinically important difference (MCID, i.e. a 1,5 point reduction) and these results can be used to benchmark MCID for the frequency and daily duration of attacks in the absence of validated ones [30,31].

Our study also provides important insights for designing clinical trials in RP. First, as evidenced in several other trials in the field we observed an important placebo response compared to run-in periods [15,27]. A previous study conducted by our team showed that regression towards the mean accounted for a large part of the placebo response in clinical trials on RP, particularly when the severity thresholds for participating in the trial were high [15]. In the present study however, we deliberately decided not to set trial eligibility thresholds in order to limit this phenomenon, and we performed repeated baseline assessments during washout periods throughout the study; visually, regression towards the mean seems weaker than in the previous series of N-of-1 trials that we had conducted, although we have not formally compared both series. The most striking finding of this work is that individual preference for one treatment over another maximizes responses to both placebo and active treatments, particularly with regard to the frequency and duration of RP attacks, thus suggesting that beyond regression towards the mean, a real and modifiable placebo effect exists in RP. While open-label (“honest”) placebos with explanations have been suggested to be beneficial in other symptomatic conditions such as chronic low back pain or irritable bowel syndrome [32], these results suggest that placebo might be a therapeutic option in RP.

An inherent limitation of studies on RP is the effect of environmental conditions, particularly temperature. We mitigated this by including in our statistical models the daily temperature recorded at the weather station closest to the patient’s home, which allowed for precise adjustment for temperature. Furthermore, the fact that we did not define severity criteria for inclusion in the trial, in order to minimize the phenomenon of regression toward the mean, may have minimized the effect of the treatment. Finally, although we trained patients to fill out their paper diaries, some were not completed correctly, with free text that was difficult to understand, resulting in a significant workload to digitize these data for analysis, and causing missing data for certain days and completely uninterpretable data for one patient. The use of a smartphone app to record Raynaud’s phenomenon attacks is a promising way to improve these aspects [33].

## Conclusion

In conclusion, our series on N-of-1 trials did not show that beetroot juice or L-citrulline are effective in improving Raynaud’s phenomenon. However, a potential reduction in the duration of attacks with beetroot juice remains to be further investigated. These findings confirm the magnitude of the placebo response in RP and suggests that patient preference is an important driver of this response.

## Data Availability

All data produced in the present study are available upon reasonable request to the authors

