## Supplementary material for "Pharmaco-nutritional strategies to increase nitric oxide signaling in Raynaud’s phenomenon (Nivose): a series of N-of-1 trials"

**Supplementary Table 1**. Non-inclusion criteria in Nivose trial.

- Uncontrolled hypertension, uncontrolled diabetes mellitus, uncontrolled angina, recent history of stroke or myocardial infarction
- Haemodynamic instability, severe hypotension (blood pressure < 90/50 mmHg)
- Severe hepatic impairment
    - Severe renal impairment (creatinine clearance < 30 ml/min)
    - Bleeding disorders
- Pregnancy (or considering pregnancy in next 4 months) or breast feeding
- Subject in an exclusion period from another study,
- subject under administrative or judicial supervision
- Subject who would receive more than 4500 euros in compensation due to his participation in other biomedical research in the 12 months preceding this study, if applicable
- Subject not able to be contacted in case of emergency

**Supplementary Figure 1.** Distribution of number of winters, cycles and days of follow-up of the 21 patients included.

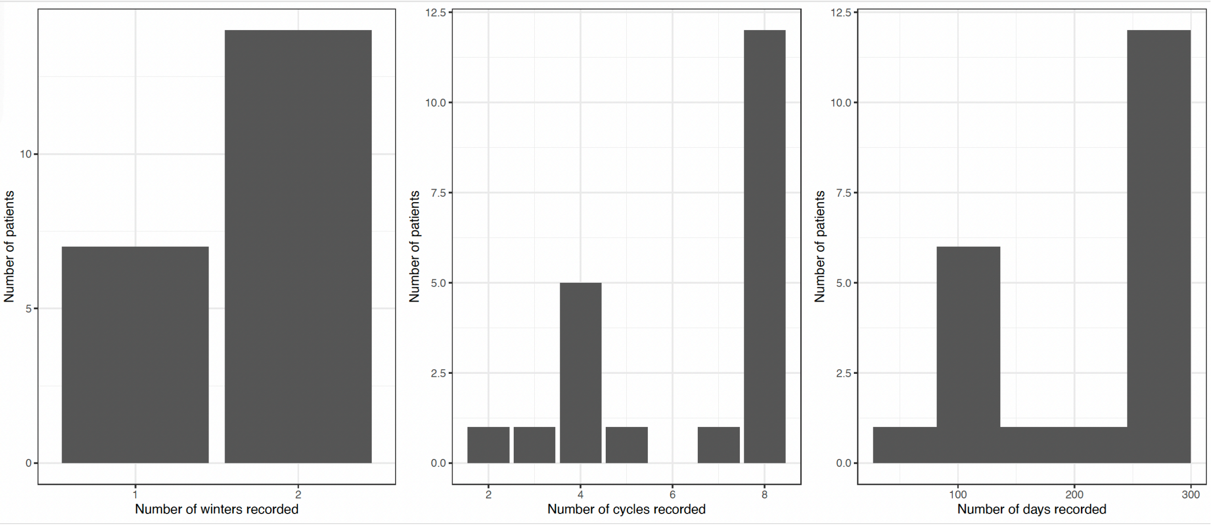

**Supplementary Table 2**. Description of baseline severity of RP in included patients, along with choice in treatment preference, main outcome and threshold for reduction.

| Patient | Treatment preference | Main outcome | Individual efficacy margin (% of reduction) | Mean RCS | Mean attacks/day | Mean duration/day |
|---|---|---|---|---|---|---|
| 1 | None | RCS | 50 | 3.55 | 1.55 | 80.00 |
| 2 | None | RCS | 50 | 3.43 | 0.62 | 23.31 |
| 3 | Beetroot | RCS | 40 | 6.29 | 1.71 | 71.79 |
| 4 | None | RCS | 30 | 3.27 | 1.13 | 33.56 |
| 5* | Beetroot | duration | 75 | 8.29 | NaN | NaN |
| 6 | Citruline | duration | 33 | 2.43 | 0.93 | 22.86 |
| 7 | None | duration | 50 | 6.75 | 1.33 | 195.42 |
| 8 | None | nattacks | 50 | 2.29 | 1.36 | 30.00 |
| 9 | Beetroot | RCS | 50 | 6.50 | 3.50 | 265.00 |
| 10 | Citruline | RCS | 20 | 2.57 | 0.64 | 16.14 |
| 11 | Beetroot | nattacks | 50 | 2.64 | 1.00 | 54.64 |
| 12 | Beetroot | nattacks | 30 | 5.64 | 2.14 | 186.00 |
| 13 | Citruline | RCS | 30 | 3.15 | 0.92 | 50.69 |
| 14 | None | duration | 50 | 3.77 | 0.79 | 62.31 |
| 15 | Beetroot | duration | 50 | 0.50 | 0.21 | 7.14 |
| 16 | None | RCS | 50 | 4.21 | 2.50 | 45.86 |
| 19 | Beetroot | RCS | 40 | 3.15 | 1.14 | 29.07 |
| 20 | Citruline | RCS | 50 | 3.79 | 1.79 | 73.93 |
| 21 | Citruline | nattacks | 50 | 1.43 | 0.36 | 10.36 |
| 22 | None | nattacks | 50 | 2.21 | 1.29 | 10.21 |
| 21 | None | duration | 75 | 6.00 | 1.00 | 29.86 |

**Supplementary Figure 2**. Squeeze plots representing severity of RP at baseline and during wash out periods.

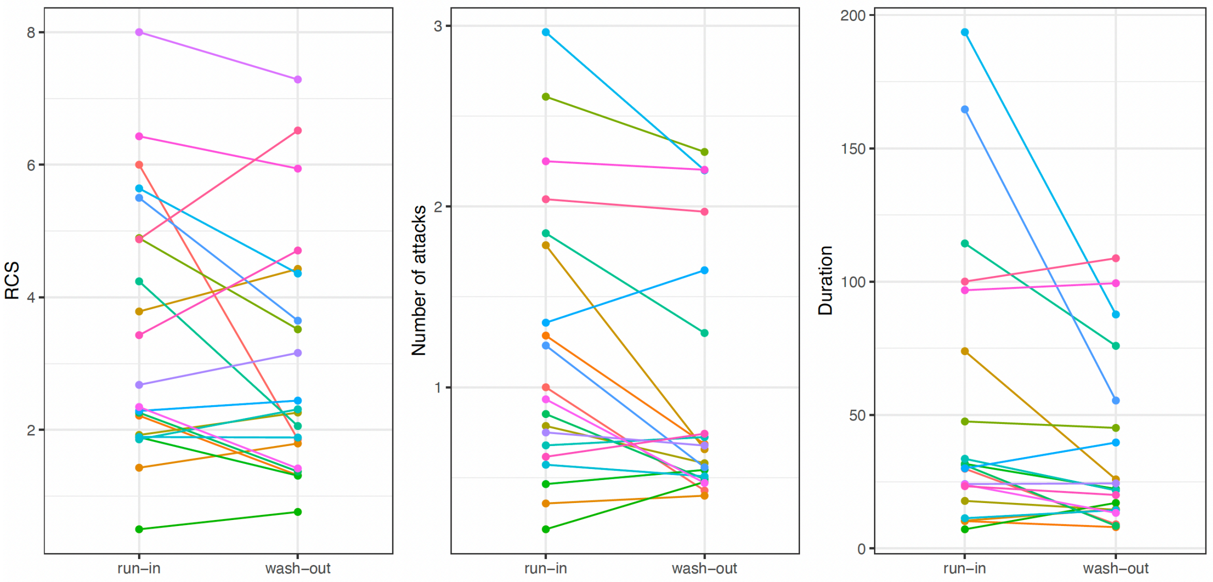
